# Polygenic risk for psychiatric disorder reveals distinct association profiles across social behaviour in the general population

**DOI:** 10.1101/2021.06.28.21259532

**Authors:** Fenja Schlag, Andrea Allegrini, Jan Buitelaar, Ellen Verhoef, Marjolein van Donkelaar, Robert Plomin, Kaili Rimfeld, Simon Edward Fisher, Beate St Pourcain

## Abstract

Many complex psychiatric disorders are characterised by a spectrum of social difficulties. These symptoms lie on a behavioural dimension that is shared with social behaviour in the general population, with substantial contributions of genetic factors. However, shared genetic links may vary across psychiatric disorders and social symptoms. Here, we systematically investigate heterogeneity in shared genetic liabilities with attention-deficit/hyperactivity disorder (ADHD), autism spectrum disorders (ASD), bipolar disorder (BP), major depression (MD) and schizophrenia, across a spectrum of different social symptoms. Specifically, longitudinally assessed low-prosociality and peer-problem scores in two UK population-based/community-based cohorts (ALSPAC, N ≤ 6174, 4-17 years; TEDS, N ≤ 7112, 4-16 years; parent- and teacher-reports) were regressed on polygenic risk scores for ADHD, ASD, BP, MD, and schizophrenia, as informed by genome-wide summary statistics from large consortia, using negative binomial regression models. Across ALSPAC and TEDS, we replicated univariate polygenic associations between social behaviour and risk for ADHD, MD, and schizophrenia. Modelling univariate genetic effects across both cohorts with random-effect meta-regression revealed evidence for polygenic links between social behaviour and ADHD, ASD, MD, and schizophrenia risk, but not BP, where differences in age, reporter and social trait captured 45-88% in univariate effect variation. For ADHD, MD, and ASD polygenic risk, we identified stronger association with peer problems than low prosociality, while schizophrenia polygenic risk was solely associated with low prosociality. The identified association profiles suggest marked differences in the social genetic architecture underlying different psychiatric disorders when investigating population-based social symptoms across 13 years of child and adolescent development.

## Introduction

Many heritable psychiatric disorders such as attention-deficit/hyperactivity disorder (ADHD), autism spectrum disorders (ASD), bipolar disorder (BP), major depression (MD) or schizophrenia are characterised by social-behavioural difficulties. In ADHD, these predominantly include peer problems^1^, while ASD is characterised by deficits in social interaction and communication^2^. Individuals with BP can suffer from social withdrawal and poor social functioning^3^, and, similarly, those with MD may show social withdrawal and disrupted social processing^4^. Individuals with schizophrenia often have poor social cognition and lack social interest^5^.

The underlying social-behavioural difficulties can be diverse. They may reflect a lack of positive interactions involving low prosocial behaviour reflected in limited helping, sharing and cooperating with others^6^. Alternatively, peer problems describe problematic interactions such as social withdrawal, being bullied, and the inability to get along with others^7^. Moreover, social symptoms change throughout development and across different social environments^8^. Therefore, social-behavioural difficulties may reflect different problems depending on developmental stage, social environment as reported by teachers or parents, and different types of skill sets. The aetiology of social problems in psychiatric disorders has, however, been little characterised.

Social behaviour is known to be heritable. Twin studies have reported heritability estimates of 0.38 to 0.76^9–11^ for prosocial behaviour and 0.41 to 0.83^9,12^ for peer problems, with variation across developmental stages^11,12^, reporters^9,10^ and social traits^9^ in community-based samples. Heritability estimates as captured by single-nucleotide polymorphisms (SNPs; SNP-h^2^) range between 0.02 to 0.27 for parent-rated peer problems in the general population, with larger estimates during adolescence compared to childhood^12^ suggesting developmental changes in genetic architectures.

One of the grand challenges in psychiatric genetics is to understand how common genetic risk, can manifest as a spectrum of diverse symptoms. Large genome-wide efforts in psychiatric genetics have demonstrated the SNP-h^2^ of ADHD (0.22)^13^, ASD (0.11)^14^, BP (0.18)^15^, MD (0.09)^16^ and schizophrenia (0.22; Supplementary Table 1)^17^. Social-behavioural difficulties in psychiatric disorder can be understood as the extreme end of a behavioural dimension that is shared with social traits in the general population^18^. These may be captured by polygenic links that differ developmentally, by different reporters and across different social symptoms, potentially reflecting distinct psychopathologies. A systematic comparison of social-behavioural difficulties, especially across neurodevelopmental disorders has, however, not yet been carried out.

In this open science framework registered study^19^, we systematically investigate genetic links between psychiatric disorders and child and adolescent social behaviour in the general population, studying the heterogeneity in polygenic associations across different ages, reporters and social traits, via a two-stage approach:

Within stage 1, we assess the relationship between polygenic risk scores for ADHD, ASD, BP, MD, and schizophrenia risk, as studied by large clinical consortia, and social-behavioural scores for low prosociality and peer problems between the ages of 7 and 17 years, as reported by parents or teachers in the population-based UK Avon Longitudinal Study for Parents and Children (ALSPAC)^20^, and follow up findings with matching scores (4-16 years; parent- and teacher-reports) in the UK community-based Twins Early Development Study (TEDS)^21^.

Within stage 2, we model differences in polygenic associations as predicted by age-, reporter-, and trait-specific social-behavioural symptoms using a random-effects meta-regression approach, combining univariate findings from ALSPAC and TEDS, and identify and compare social-behavioural association patterns for each disorder.

## Samples and methods

### Genome-wide summary statistics for psychiatric disorder

We studied genome-wide summary statistics for five psychiatric disorders as published by the Psychiatric Genomic Consortium (PGC), the Danish Lundbeck Foundation Initiative for Integrative Psychiatric Research (iPSYCH) and/or the UK Biobank (UKBB): ADHD-PGC/iPSYCH^13^, ASD-PGC/iPSYCH^14^, BP-PGC^15^, MD-PGC/UKBB^16^, and schizophrenia-PGC^17^. Cohort details including ancestry, size, imputation reference panel, symptoms and age-of-onset of the disorder are described in the Supplement (Supplementary Information, Supplementary Table 1).

### Social behaviour in the general population

<u>ALSPAC</u> is a UK population-based longitudinal pregnancy-ascertained birth cohort with birth dates between 1991 and 1992^20,22^. Ethical approval for the study was obtained from the ALSPAC Ethics and Law Committee and the Local Research Ethics Committees. Consent for biological samples has been collected in accordance with the Human Tissue Act (2004). Informed consent for the use of data collected via questionnaires and clinics was obtained from participants following the recommendations of the ALSPAC Ethics and Law Committee at the time (Supplementary Information).

<u>TEDS</u> is a community-based longitudinal study of >10,000 twin pairs representative of England and Wales, recruited from 1994 to 1996 births^21^. Ethical approval for the study was granted by King’s College London’s ethics committee for the Institute of Psychiatry, Psychology and Neuroscience (05.Q0706/228), and written informed consent was given by the parents prior to data collection.

#### Phenotype information

Prosocial behaviour and peer problems were assessed in ALSPAC and TEDS children (Supplementary Information, Table 1) using standardised behavioural screening questionnaires. Both, prosocial behaviour (here recoded as low-prosociality scores) and peer problems were assessed using subscales of the Strengths-and-Difficulties questionnaire (SDQ^23^), based on parent- and teacher-reports at the same ages. In ALSPAC, parent-reported (predominantly mother-reported) behaviour was measured at the ages of 7, 10, 12, 13 and 17 years and in TEDS at the ages of 4, 7, 9, 11, 14, and 16 (prosocial scores only) years. In addition, teacher reports were obtained at the ages of 8 and 11 years in ALSPAC and at the ages of 9, 12, and 14 years in TEDS. Both scores are phenotypically modestly to moderately correlated with each other (Supplementary Tables 2,3).

**Table 1:** Descriptive information of low prosociality and peer problems in ALSPAC and TEDS.

|  |  | Age (years) | Variable score | % Males | N |
| --- | --- | --- | --- | --- | --- |
|  |  | Mean (SD) | Mean (SD) |  |  |
| ALSPAC |  |  |  |  |  |
| Low prosociality<sup>1</sup> |  |  |  |  |  |
| Parent-reported: | 7Y | 6.79 (0.11) | 1.82 (1.75) | 51 | 5610 |
|  | 10Y | 9.65 (0.12) | 1.66 (1.65) | 50 | 5670 |
|  | 12Y | 11.72 (0.13) | 1.65 (1.68) | 50 | 5268 |
|  | 13Y | 13.16 (0.18) | 2.76 (1.73) | 50 | 5069 |
|  | 17Y | 16.84 (0.36) | 1.97 (1.87) | 48 | 4151 |
| Teacher-reported: | 8Y | 8.33 (0.31) | 2.21 (2.42) | 50 | 3686 |
|  | 11Y | 11.16 (0.33) | 2.06 (2.35) | 50 | 4417 |
| Peer problems |  |  |  |  |  |
| Parent-reported: | 7Y | 6.79 (0.11) | 1.02 (1.04) | 51 | 5608 |
|  | 10Y | 9.65 (0.12) | 1.1 (1.49) | 50 | 5661 |
|  | 12Y | 11.72 (0.13) | 1.1 (1.56) | 50 | 5263 |
|  | 13Y | 13.16 (0.18) | 1.19 (1.61) | 50 | 5061 |
|  | 17Y | 16.84 (0.36) | 1.11 (1.51) | 48 | 4156 |
| Teacher-reported: | 8Y | 8.33 (0.31) | 1.13 (1.74) | 50 | 3689 |
|  | 11Y | 11.16 (0.33) | 1.2 (1.85) | 50 | 4417 |
| TEDS |  |  |  |  |  |
| Low prosociality<sup>1</sup> |  |  |  |  |  |
| Parent-reported: | 4Y | 4.04 (0.12) | 2.6 (1.86) | 48 | 6958 |
|  | 7Y | 7.06 (0.25) | 1.84 (1.79) | 48 | 7112 |
|  | 9Y | 9.01 (0.29) | 2.71 (1.71) | 47 | 3375 |
|  | 11Y | 11.25 (0.7) | 1.46 (1.65) | 48 | 6039 |
|  | 16Y | 16.31 (0.68) | 1.74 (1.94) | 45 | 5252 |
| Teacher-reported: | 7Y | 7.2 (0.28) | 2.68 (2.36) | 49 | 5900 |
|  | 9Y | 9.03 (0.29) | 2.44 (2.26) | 47 | 2825 |
|  | 12Y | 11.5 (0.66) | 1.99 (2.09) | 47 | 4931 |
| Peer problems |  |  |  |  |  |
| Parent-reported: | 4Y | 4.04 (0.12) | 1.52 (1.54) | 48 | 6948 |
|  | 7Y | 7.06 (0.25) | 1.01 (1.45) | 48 | 7112 |
|  | 9Y | 9.01 (0.29) | 1.11 (1.59) | 47 | 3370 |
|  | 11Y | 11.25 (0.7) | 1.11 (1.54) | 48 | 6023 |
| Teacher-reported: | 7Y | 7.2 (0.28) | 1.07 (1.48) | 49 | 5900 |
|  | 9Y | 9.03 (0.29) | 0.85 (1.47) | 47 | 2828 |
|  | 12Y | 11.51 (0.66) | 1.04 (1.6) | 47 | 4964 |

<sup>1</sup> Reverse coded SDQ prosocial scale.
ALSPAC - Avon Longitudinal study of Parents and Children; SDQ - Strengths-and-Difficulties questionnaire; TEDS - Twins Early Development Study; Y - Age in years
All low-prosociality and peer-problem scores were assessed using the Strengths-and-Difficulties questionnaire.

### Univariate polygenic scoring analyses in ALSPAC and TEDS

#### Polygenic scoring analyses

Consistent with current guidelines^24^, we constructed polygenic risk scores (PRS) for each disorder (ADHD, ASD, BP, MD and schizophrenia) within ALSPAC and TED using a clumping and thresholding approach (risk-variant selection thresholds 0.001 ≤ *P*_T_ <1), based on high-quality genome-wide imputed SNPs (Supplementary Information).

Within ALSPAC, we studied unrelated children and adolescents (genomic relatedness < 0.125). We regressed untransformed social-behavioural scores (peer problem or low prosociality) on Z-standardised PRS using a negative binomial model, adjusting for covariate effects of sex, age, and the first two PCs (R:MASS; Supplementary Information). Within TEDS, we analysed pairs of dizygotic twins and a single twin of each monozygotic pair. PRS association analyses were carried out using a multi-level negative binomial regression approach (R:lme4, v.1.1-26^25^), with a random intercept to adjust for family relatedness, and fixed effects for PRS adjusting for covariate effects of sex, age, the first ten PCs, genotyping-batch, and genotyping-chip effects. For both models the negative binomial and multi-level negative binomial model, beta coefficients indicate the change in log counts of social score by one SD change in PRS (PRS effects). We tested the predictive ability of PRS using ΔMcFadden’s-R^2^(Supplementary Information)^26^.

#### Multiple-testing correction

Using Matrix Spectral Decomposition (matSpD)^27^, we accounted for 14 interrelated social-behavioural scores (Supplementary Table 2) and five psychiatric disorders in ALSPAC by identifying an effective number of 10 independent variables, and adjusting the multiple-testing burden of all univariate PRS analyses to 0.05/(10*5)=0.001. For follow-up analyses in TEDS, with an effective number of 12 independent variables, the multiple-testing burden under a one-sided test was adjusted to 0.1/(12*5)=0.0017, accounting for 15 interrelated scores and five psychiatric disorders (Supplementary Table 3).

#### Power analyses

We estimated the power to detect PRS effects in the discovery cohort (ALSPAC) using the R software package avengeme^28^ (Supplementary Information), assessing the influence of trait- or disorder-specific SNP-h^2^ and trait-disorder covariance.

### Meta-regression of polygenic effects

#### Meta-regression models

For each disorder, we combined univariate PRS estimates for SDQ subscales across ALSPAC and TEDS using a random-effect meta-regression model (R:metafor, v.2.1-0^29^, Supplementary Information). In brief, we systematically assessed whether heterogeneity in PRS association effects (based for simplicity on a representative PRS risk variant selection threshold of *P*_T_ ≤ 0.1) can be attributed to differences in social behaviour explained by the median age of assessment, reporter (parent vs teacher), and SDQ-based social trait (low prosociality versus peer problems). For each disorder, we fitted a full model including a random intercept accounting for repeated measures (nested within cohort) as well as fixed effects for age-, reporter-, trait- and cohort-specific effects. The most parsimonious model was identified by dropping successively fixed effects from the model (likelihood-ratio test at *P* > 0.05) and assessing residual heterogeneity (Cochran’s Q test, Supplementary Information). The inter-relatedness of PRS association effects across SDQ-based social scores within each cohort was accounted for by constructing a composite variance covariance matrix analogous to models accounting for correlated phylogenetic histories^30^.

#### Multiple-testing correction

A threshold of *P* ≤ 0.01 (0.05/five disorders) was applied.

## Results

### Stage 1: Univariate association analyses

#### Discovery analyses in ALSPAC

We assessed the univariate association between each of the 14 population-based social-behavioural scores in ALSPAC, including low-prosociality and peer problem scores between the ages of 7 and 17 years, as reported by parents or teachers, and five disorder-specific PRS (Supplementary Table 1, ADHD, ASD, BP, MD, and schizophrenia; 70 analyses; multiple-testing threshold of *P* ≤ 0.001). All social scores were skewed, with most children showing little difficulties in prosocial behaviour and peer interactions (Table 1). Consequently, we assessed genetic associations with a negative binomial regression model, given the better fit of a count data model compared to a linear model (Supplementary Table 4). PRS effects were fitted across multiple risk variant selection thresholds (0.001 ≤ *P*_T_ <1, Supplementary Tables 5,6, Figure 1), but are here, for simplicity, reported at *P*_T_ ≤ 0.1.

**Figure 1:**
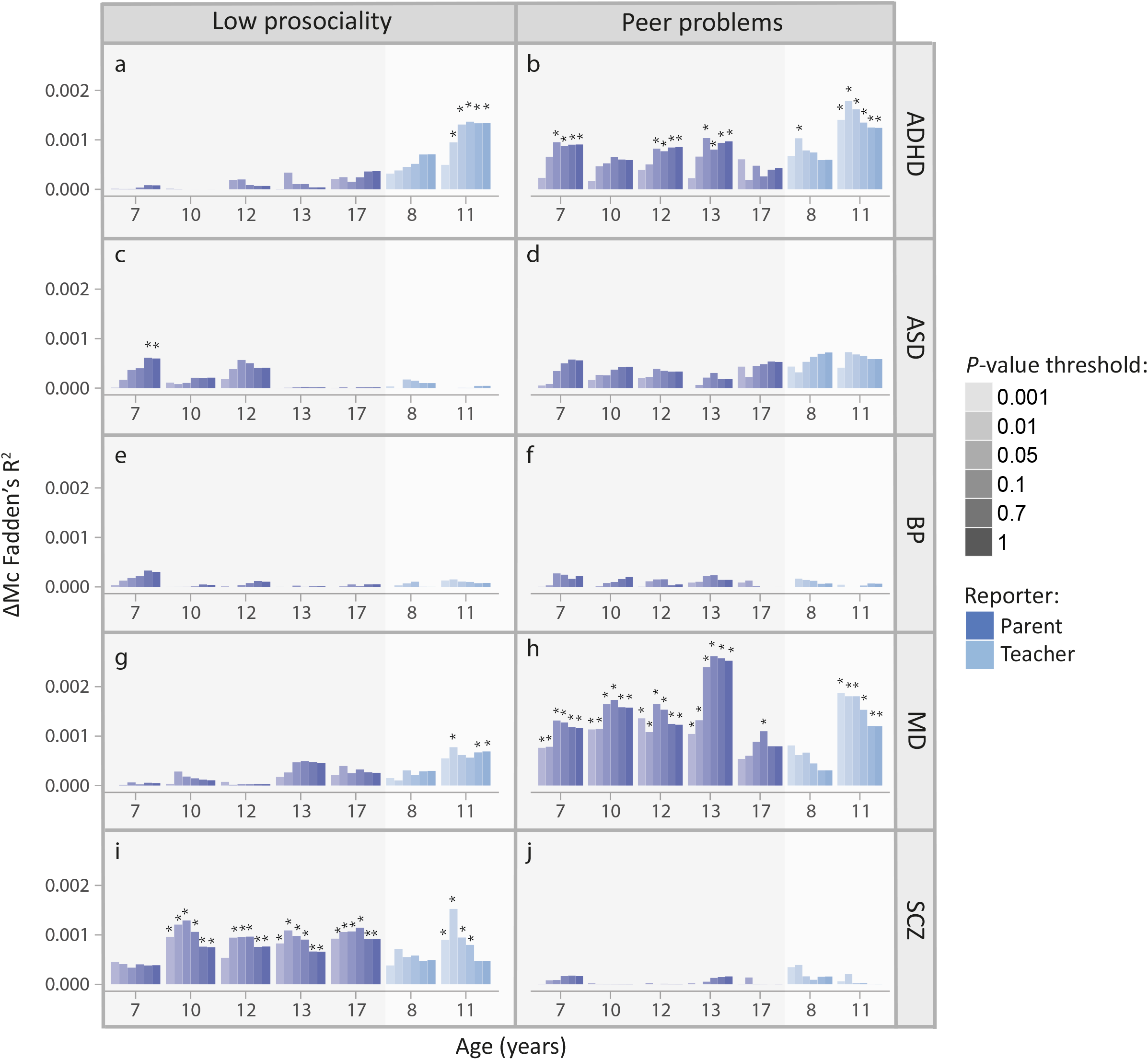
Polygenic scoring analyses between social behaviour and psychiatric disorder in ALSPAC. ΔMcFadden’s-R^2^ is shown for the prediction of low-prosociality and peer-problem scores by ADHD-PRS (a, b), ASD-PRS (c, d), BP-PRS (e,f), MD-PRS (g, h), SCZ-PRS (i, j). Psychiatric disorder samples (ADHD-PGC/iPSYCH, ASD-PGC/iPSYCH, BP-PGC, MD-PGC/UKBB, and SCZ-PGC) were used to construct Z-standardised PRS in ALSPAC (ADHD-PRS, ASD-PRS, BP-PRS, MD-PRS, and SCZ-PRS) at multiple *P*-value thresholds. Association analyses with social behaviour (low prosociality and peer problems) were conducted using negative binomial regression (multiple-testing corrected *P*-value: *P≤0.001). ADHD - Attention-deficit/hyperactivity disorder; ALSPAC - Avon Longitudinal study of Parents and Children; ASD - Autism spectrum disorders; BP - Bipolar disorder; iPSYCH - Lundbeck Foundation Initiative for Integrative Psychiatric Research; MD- Major depression; PGC - Psychiatric Genomics consortium; PRS-Polygenic risk scores; *P*_T_ - PRS threshold; SCZ - Schizophrenia Low-prosociality and peer-problem scores were assessed using the Strengths-and-Difficulties questionnaire.

Many social-behavioural scores were associated with polygenic risk for ADHD, MD and schizophrenia. For ADHD-PRS, the strongest association was identified for teacher-reported peer problems at the age of 11 years (β_ADHD_11Y_(SE)=0.10(0.025), ΔMcFadden’s-R^2^=0.0013, *P*=2.5×10^−5^; Figure1a,b). MD-PRS was most strongly associated with parent-reported peer problems scores at the age of 13 years (β_MD_13Y_(SE)=0.12(0.019), ΔMcFadden’s-R^2^=0.0026, *P*=2.6×10^−10^; Figure1g,h). Associations between schizophrenia-PRS and social traits were strongest for teacher-rated low-prosociality scores at 11 years (β_SCZ_11Y_(SE)=0.07(0.019), ΔMcFadden’s-R^2^=8.0×10^−4^, *P*=2.2×10^−4^; Figure1i,j). For ASD-PRS, no univariate association with social symptoms at *P*_T_ ≤ 0.1 passed the multiple-testing threshold. However, at less stringent *P*_T_ thresholds, association with parent-reported low prosociality at the age of 7 years was present (for example at *P*_T_ < 0.5, β_ASD_7Y_(SE)=0.045(0.013), ΔMcFadden’s-R^2^=5.8×10^−4^, *P*=6.6×10^−4^; Figure1c,d) and meta-analysing univariate ASD-PRS effects across *P*_T_ ≤ 0.1 yielded further support for association (data not shown). There was little evidence for association between BP-PRS and any of the studied social measures (Figure1e,f).

Power analyses showed that across all studied psychiatric conditions, our study had sufficient power under the assumption of fixed trait-disorder covariance (equivalent to the SNP-h^2^ of the disorder; Supplementary Table 7; Supplementary Information; Supplementary Figures 1,2; dotted lines). Once data-driven covariance patterns and, thus, trait architectures were considered, power curves followed observed associations (Supplementary Figures 1,2; solid lines), largely, independent of trait SNP-h^2^ (Supplementary Table 8). Only for polygenic BP and polygenic ASD risk (except for social scores at age 7 years) the power was <80% (at *P*_T_ < 0.1), irrespective of the studied social symptoms, consistent with the lack in univariate associations.

#### Follow-up analyses in TEDS

Subsequently, we studied the univariate association of polygenic risk for ADHD, ASD, BP, MD, and schizophrenia (at 0.001 ≤ *P*_T_ < 1) with 15 ALSPAC-matching community-based social-behavioural scores in TEDS. Parent- and teacher-reported low-prosociality and peer-problem scores were longitudinally assessed between 4 and 16 years, showing skewed distributions (Table 1, Supplementary Table 9). At *P*_T_ ≤ 0.1, we replicated evidence for association between social-behavioural scores and polygenic risk for ADHD, MD and schizophrenia (75 analyses; Figure 2, Supplementary Tables 10,11; multiple-testing threshold of *P* ≤ 0.0017). In addition, we observed evidence for association between ASD-PRS and peer problems that was strongest in parent-reported scores at the age of 11 years (β_ASD_11Y_(SE)=0.093(0.018), ΔMcFadden’s-R^2^= 0.0015, *P*=2.7×10^−7^; Figure2g,h). As observed in ALSPAC, we did not find association between BP-PRS and any of the studied social measures in TEDS (Figure2e,f).

**Figure 2:**
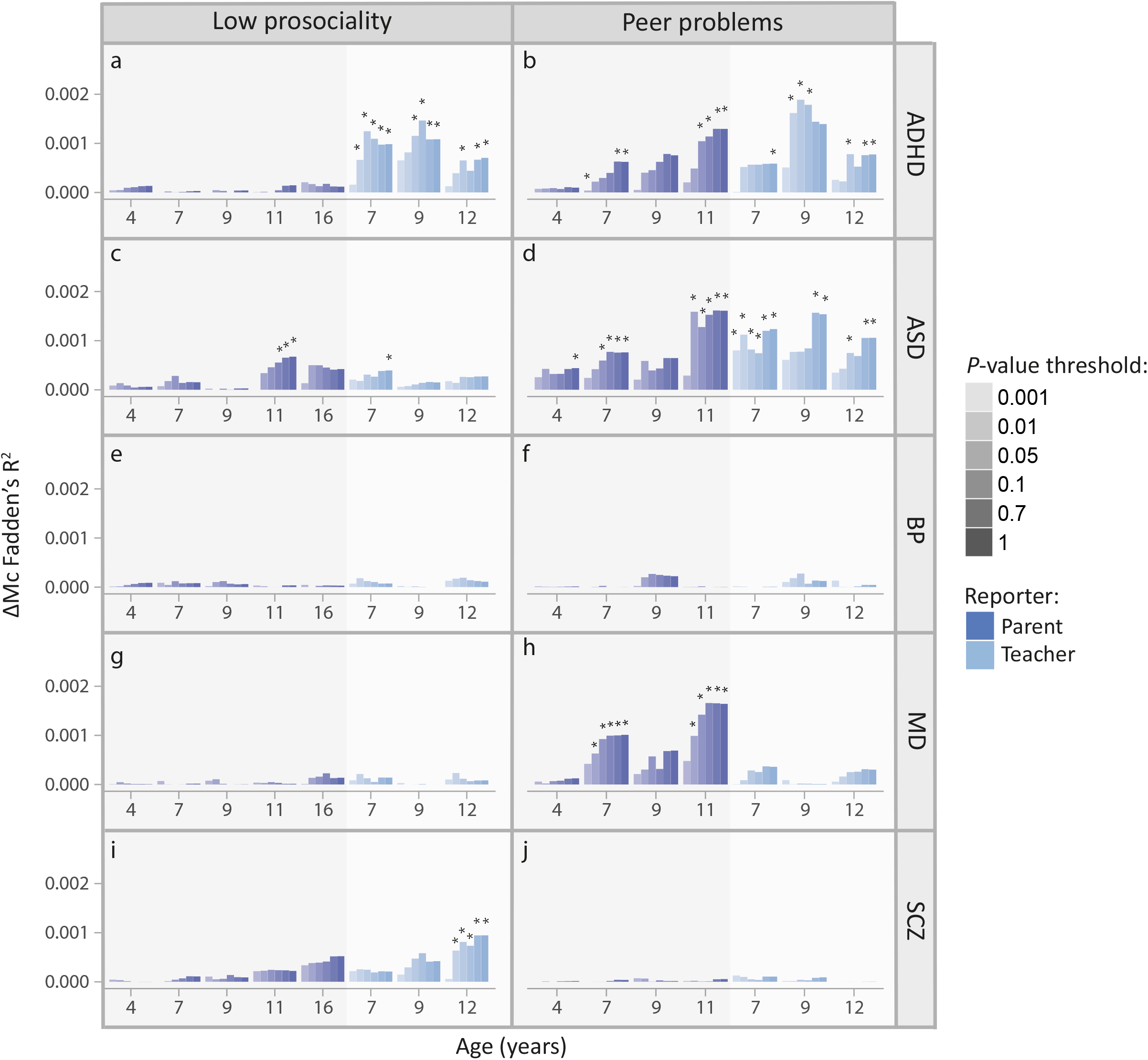
Polygenic scoring analyses between social behaviour and psychiatric disorder in TEDS. ΔMcFadden’s-R^2^ is shown for the prediction of low-prosociality and peer-problem scores by ADHD-PRS (a, b), ASD-PRS (c, d), BP-PRS (e,f), MD-PRS (g, h), SCZ-PRS (i, j). Psychiatric disorder samples (ADHD-PGC/iPSYCH, ASD-PGC/iPSYCH, BP-PGC, MD-PGC/UKBB, and SCZ-PGC) were used to construct Z-standardised PRS in TEDS (ADHD-PRS, ASD-PRS, BP-PRS, MD-PRS, and SCZ-PRS) at multiple *P*-value thresholds. Association analyses with social behaviour (low prosociality and peer problems) were conducted using negative binomial regression (multiple-testing corrected one-sided *P*-value: \**P*≤0.0017). ADHD - Attention-deficit/hyperactivity disorder; ASD - Autism spectrum disorders; BP - Bipolar disorder; iPSYCH - Lundbeck Foundation Initiative for Integrative Psychiatric Research; MD-Major depression; PGC - Psychiatric Genomics consortium; PRS-Polygenic risk scores; *P*_T_ - PRS threshold; SCZ – Schizophrenia; TEDS - Twins Early Development Study Low-prosociality and peer-problem scores were assessed using the Strengths-and-Difficulties questionnaire.

### Stage 2: Meta-regression of polygenic association signals in ALSPAC and TEDS

For each disorder, we combined univariate polygenic PRS estimates for the 29 SDQ-based social scores from both ALSPAC and TEDS, using a random-effects meta-regression approach (5 analyses, multiple-testing threshold *P*≤0.01). Specifically, we modelled heterogeneity in PRS effect estimates as predicted by age-, reporter-, and trait-specific differences in social behaviour, captured by the fixed-effect meta-regression estimates 0. For each disorder, we first fitted a full meta-regression model and, subsequently, dropped predictors to identify the most parsimonious model based on likelihood-ratio tests (Supplementary Tables 12,13, Supplementary Figures 3-7).

Meta-regression analyses provided evidence for association between social behaviour and PRS for ADHD, ASD, MD, and schizophrenia, but not BP. Across disorders, polygenic effects varied with age, reporter, and, especially, social trait (Table 2). As there was little evidence for cohort-specific fixed effects, these effects were omitted from the most parsimonious models throughout. For ADHD-PRS, the most parsimonious meta-regression model provided evidence for an increase in PRS effects with age (*θ*_age(Y)_(SE)=0.0025(8.9×10^−4^), *P*=0.0042), teacher-reported scores (*θ*_teacher_report_(SE)=0.044(0.0085), *P*=2.5×10^−7^) and peer problems (*θ*_peer_problems_(SE)=0.03(0.0089), *P*=7.3×10^−4^). Likewise, the meta-regression model for MD-PRS showed an increase in PRS effect with age (*θ*_age(Y)_(SE)= 0.0035(9.5×10^−4^), *P*=1.9×10^−4^) and peer problems (*θ*_peer_problems_(SE)=0.048(0.0093), *P*=2.8×10^−7^). In contrast to ADHD and MD, the most parsimonious model for schizophrenia revealed a decrease in PRS effects for peer problems (*θ*_peer_problems_(SE)=-0.027(0.0094), *P*=0.0033). As there was a trend for a small positive age-effect that captured a considerable proportion of effect heterogeneity, this effect was retained in the model. For ASD-PRS, we observed an increase in PRS effect for peer problems (*θ*_peer_problems_(SE)=0.037(0.0083), *P*=7.9×10^−6^) consistent with meta-analytic but not univariate analyses in ALSPAC. The most parsimonious model based on BP-PRS revealed little evidence for association with any social symptoms.

**Table 2:** Random-effects meta-regression across psychiatric PRS effects on social-behavioural symptoms^1^.

| **ADHD-PRS ($R^2=0.88$)** |  |  |  |
| --- | --- | --- | --- |
| **Parameter** | **$\theta$ (SE)** | **Z-value** | **P-value** |
| Intercept (Age 4, parent-reported, low prosociality) | -0.015 (0.01) | -1.49 | 0.14 |
| Age (Centered at 4 years) | 0.0025 (0.00089) | 2.86 | 0.0042 |
| Reporter (Teacher-reported) | 0.044 (0.0085) | 5.16 | $2.5 \times 10^{-7}$ |
| Trait (Peer problems) | 0.03 (0.0089) | 3.38 | $7.3 \times 10^{-4}$ |
| **ASD-PRS ($R^2=0.58$)** |  |  |  |
| **Parameter** | **$\theta$ (SE)** | **Z-value** | **P-value** |
| Intercept (Low prosociality) | 0.021 (0.0063) | 3.36 | $7.7 \times 10^{-4}$ |
| Trait | 0.037 (0.0083) | 4.47 | $7.9 \times 10^{-6}$ |
| **BP-PRS ($R^2=0.00$)** |  |  |  |
| **Parameter** | **$\theta$ (SE)** | **Z-value** | **P-value** |
| Intercept | 0.0054 (0.0056) | 0.98 | 0.33 |
| **MD-PRS ($R^2=0.84$)** |  |  |  |
| **Parameter** | **$\theta$ (SE)** | **Z-value** | **P-value** |
| Intercept (Age 4, low prosociality) | -0.018 (0.011) | -1.66 | 0.096 |
| Age (Centered at 4 years) | 0.0035 (0.00095) | 3.74 | $1.9 \times 10^{-4}$ |
| Trait (Peer problems) | 0.048 (0.0093) | 5.14 | $2.8 \times 10^{-7}$ |
| **Schizophrenia-PRS ($R^2=0.45$)** |  |  |  |
| **Parameter** | **$\theta$ (SE)** | **Z-value** | **P-value** |
| Intercept (Low prosociality) | 0.017 (0.011) | 1.55 | 0.12 |
| Age (Centered at 4 years) | 0.0018 (0.00096) | 1.86 | 0.063 |
| Trait (Peer problems) | -0.027 (0.0094) | -2.94 | 0.0033 |

<sup>1</sup> PRS association effects for ADHD, ASD, BP, MD and schizophrenia risk (based on negative binomial regression) were combined across 29 social symptoms (14 ALSPAC-based + 15 TEDS-based; at $P_T \leq 0.1$ ) using random-effects meta-regressions, accounting for phenotypic correlations between social scores. Here, the most parsimonious models are shown with the predictors ( $\theta$ ) of PRS effect heterogeneity including age-, reporter- (parent versus teacher reports), and trait-specific differences in social behaviour (low prosociality versus peer problems).
ADHD - Attention-Deficit/Hyperactivity Disorder; ALSPAC - Avon Longitudinal study of Parents and Children; ASD - Autism spectrum disorder; BP - Bipolar disorder; MD- Major depression; PRS-Polygenic risk scores; $P_T$ - PRS threshold; TEDS - Twins Early Development Study

Predicted heterogeneity in PRS effects for ADHD, ASD, MD and (Figure 3; Supplementary Figures 3-7) can be summarised as follows: Meta-analytically predicted PRS effects 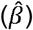 indicate an association of ADHD-PRS with low prosociality based on teacher-reports 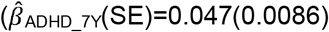 to 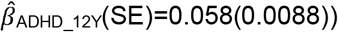 and for parent-reports only from the age of 11 years onwards 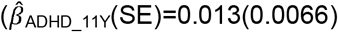 to 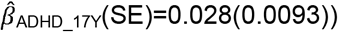; ADHD-PRS are also associated with peer problems based on both parent-reports 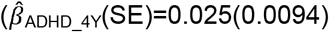 to 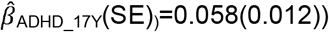 and teacher-reports 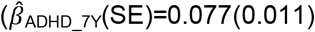 to 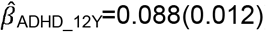), but not parent-reported low prosociality between the ages of 4 and 10 years 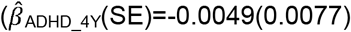 to 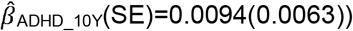. Polygenic association with MD-PRS increased with age and was larger for peer problems 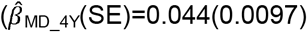 to 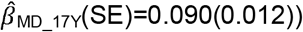 than low prosociality 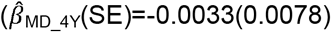 to 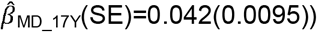 with evidence for an association with low prosociality only from the age of 9 years onwards 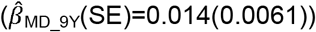. In contrast, association effects of schizophrenia-PRS risk with social behaviour were only found for low prosociality 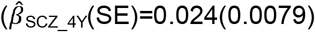 to 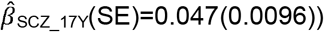, but not peer problems 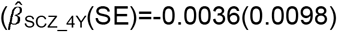 to 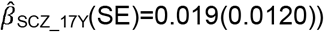. ASD-PRS association effects were stable across age, but larger for peer problems 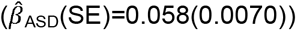 than low prosociality 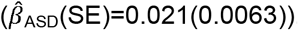.

**Figure 3:**
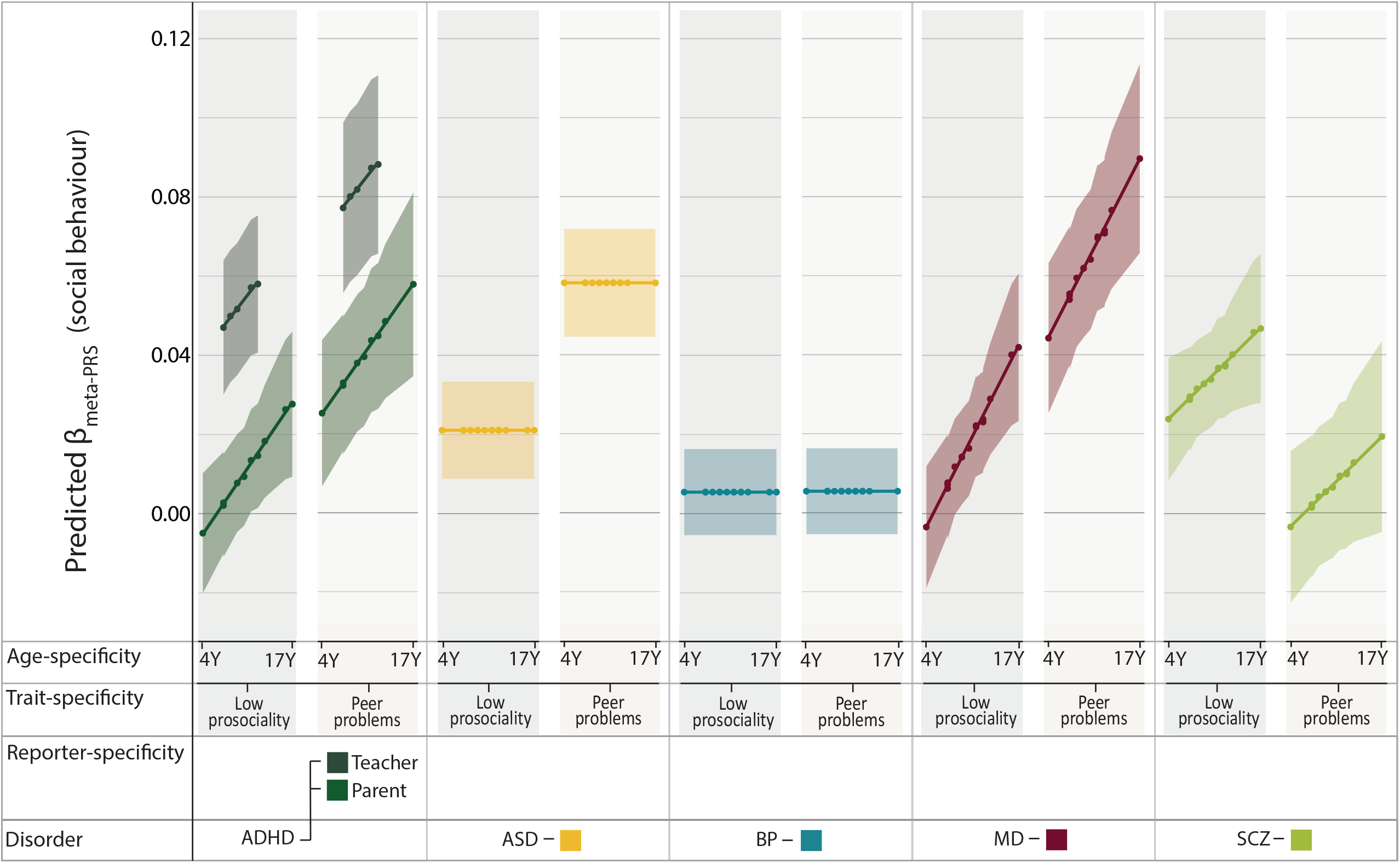
Meta-analytically predicted PRS effects for psychiatric disorder with social behaviour. For each disorder (ADHD, ASD, BP, MD and schizophrenia) 29 SDQ-based PRS effects from ALSPAC and TEDS (at *P*_T_ ≤ 0.1) were combined using random-effects meta-regression and predicted by age-, reporter- (parent versus teacher), and trait- (low prosociality versus peer problems) specific social symptoms. Predicted estimates for PRS effects (used to fit the meta-regression model) are shown as dots. ADHD - Attention-Deficit/Hyperactivity Disorder; ALSPAC - Avon Longitudinal study of Parents and Children; ASD - Autism spectrum disorders; BP - Bipolar disorder; MD- Major depression; PRS-Polygenic risk scores; *P*_T_ - PRS threshold; SCZ - Schizophrenia; SDQ - Strengths-and-Difficulties questionnaire; TEDS - Twins Early Development Study; Y - years

Together, these findings are consistent with distinct association profiles for social behaviour across psychiatric conditions.

## Discussion

Investigating polygenic links between risk for psychiatric disorder and population-based social behaviour, this study identified marked differences in genetic associations between psychiatric disorders and a spectrum of social-behavioural difficulties. We observed robust evidence for shared genetic influences between child and adolescent social difficulties and polygenic risk for ADHD, MD and schizophrenia across two large UK population-based cohorts in a univariate association approach. Combining univariate findings in a meta-regression approach, we identified further evidence for association between ASD risk and social difficulties. Here, we showed that the identified meta-analytic association profiles systematically varied with age-, reporter- and trait-specific social symptoms across disorders. These findings suggest a diverse genetic landscape of social phenotypes that is differentially shared with psychiatric disorder. As such, our results refine previous research demonstrating genetic overlap with psychiatric risk for social behaviour-related traits such as emotion recognition in childhood and adolescence^31,32^, self-reported empathy^33^, loneliness^34^, and sociability^35^ in adults.

We observed age-specific effects for ADHD, MD and schizophrenia risk indicating polygenic links with social behaviour that increased from childhood to adolescence, detectable at 4 years of age onwards. These findings confirm previously reported developmental changes in the genetic overlap of schizophrenia risk with social communication^36^. While a developmental increase in genetic association effects is in line with the typical onset of MD and schizophrenia during adolescence and adult life^37,38^, our findings may link to subthreshold social difficulties preceding clinical diagnosis^39,40^ or early-onset cases, which are thought to convey more severe symptoms^41,42^. The age-specific increase in association between risk for ADHD, a typical childhood-onset disorder, and scores for both low prosociality and peer problems during the course of development suggests that the genetic link with social behaviour may progress into adulthood^43^.

For ADHD risk only, we identified reporter-specific genetic effects, with stronger genetic links for teacher-reported compared to parent-reported social behaviour. Social behaviour at school as reported by teachers evaluates rule-oriented behaviour^44^, but also adequate peer-peer interactions among children of the same age. Therefore, school environments may, specifically, expose behavioural difficulties of children with ADHD. Problems may arise due to children’s high levels of distractibility but also their disruptive/oppositional behaviours, consistent with teacher-reported, but not parent-reported oppositional defiant disorder symptoms predicting less prosocial behaviour^1^.

Across disorders, we found evidence for social trait-specific effects shaping polygenic links with ADHD, ASD, MD and schizophrenia risk. The sole association of schizophrenia risk with low prosociality, but not peer problems, may reflect specific impairments in social cognition and a lack of social interest and empathy in psychotic disorders^5^. In contrast, the stronger genetic association of ADHD, ASD, and MD risk with peer problems, compared to prosocial scores, may reflect genetic links with socially disruptive behaviours and poor social skills, potentially related to difficulties with communication, emotion regulation, executive functioning, and/or social isolation^4,45,46^. The similarity in polygenic trait effects for ADHD, ASD, and MD is in line with previously reported similarities in social symptoms at the phenotypic level^47–49^ and genetic correlations between these conditions^50^.

The absence of genetic interrelationships with BP is consistent with previous reports^33,35,51^ and either reflects lack of power or suggests that, genetically, social symptoms may not be directly involved in the genetic aetiology of the disorder.

Given the robustness of univariate polygenic signals across two population-based/community-based cohorts, our findings suggest that the identified association profiles capture differences in genetic covariance between social traits and disorders, reflecting differences in underlying aetiological mechanisms: (1) The discordant social trait-specific association pattern for schizophrenia compared to ADHD, ASD, and MD risk suggests that the aetiological spectrum of social difficulties is distinct across disorders, supporting targeted treatment strategies for psychotic versus non-psychotic disorders^52^. (2) Teacher-specific effects shaping polygenic links with ADHD risk may reflect a subtype of social-behavioural problems within school environments that is influenced by genetic factors unique to ADHD, highlighting the importance of school-related interventions. A lack of power to detect reporter-specific effects for other disorders is a less likely explanation, despite known bias affecting parent-reported measures^53^, given association of parent-reported measures with other psychiatric PRS (Supplementary Figures 1-2). (3) Age-specific changes in genetic associations involving ADHD, MD, and schizophrenia risk, but not ASD risk, are likely to reflect the change in overlap between the largely stable symptom spectrum in disorder and developmentally highly variable social problems in the general population. These findings suggest that social-behavioural symptoms during later adolescence most strongly contribute to ADHD, MD, and schizophrenia genetic architectures underscoring the need for early-life interventions. Conversely, the lack of age-specific changes in the association of ASD risk with social behaviour suggests that these polygenic links involve social problems that already emerge before or at the age of 4 years and remain developmentally stable, consistent with the early social core deficits in ASD^2^. These findings contrast with the developmental decline in the genetic overlap of ASD risk with social communication that was previously reported^36^, possibly reflecting differences in social-behavioural versus - communication-related skills, where the latter rely more strongly on social cognition, and verbal and non-verbal communication^7^.

Our study has several strengths and limitations: We investigated two social traits as reported by parents and teachers across 13 years of child and adolescent development, studying polygenic links with multiple psychiatric conditions, using a count data approach. We robustly identified evidence for genetic association in two large UK population-based/community-based cohorts, and systematically modelled heterogeneity in polygenic estimates using a random-effects meta-regression approach. However, consistent with other PRS analyses^36^, effect sizes were small, and due to different sets of risk-increasing alleles analysed, a direct comparison of PRS effect size across disorders is not meaningful here. We exclusively investigated social symptoms with the SDQ. Different instruments, including those assessing reciprocal social interactions, might capture additional symptoms and broaden the interpretation of social difficulties in psychiatric disorders, in particular for ASD risk. Furthermore, polygenic signals might be biased by population-based phenomena, such as dynastic effects and non-random mating^54^, but also non-random missingness linked to socio-economic status^55,56^. However, this is less likely as bias would similarly affect uniformly ascertained SDQ scores, resulting in homogeneous and not heterogeneous genetic association profiles. Finally, misclassification of psychiatric disorder and phenotypic heterogeneity may increase genetic correlations across a spectrum of social symptoms^57^. Thus, further studies should refine our findings by replicating across broadly defined social phenotypes in European and non-European cohorts to promote the translation into precision medicine^52^.

In conclusion, our findings reveal marked differences in the social genetic architecture underlying different psychiatric disorders and demonstrate that social symptoms represent a heterogeneous spectrum of related endophenotypes.

## Data Availability

Data is available via the Avon Longitudinal Study for Parents and Children (ALSPAC;http://www.bristol.ac.uk/alspac/researchers/access/)and via the Twins Early Development Study (TEDS;https://www.teds.ac.uk/researchers/teds-data-access-policy). Publicly available psychiatrich summary statistics are avialable via http://www.med.unc.edu/pgc.

## Acknowledgements

We are extremely grateful to all the families who took part in this study, the midwives for their help in recruiting them, and the whole ALSPAC team, which includes interviewers, computer and laboratory technicians, clerical workers, research scientists, volunteers, managers, receptionists and nurses. The UK Medical Research Council and Wellcome (Grant ref: 217065/Z/19/Z) and the University of Bristol provide core support for ALSPAC. This publication is the work of the authors and FS and BSTP will serve as guarantors for the contents of this paper. A comprehensive list of grants funding is available on the ALSPAC website (http://www.bristol.ac.uk/alspac/external/documents/grant-acknowledgements.pdf).

GWAS data was generated by Sample Logistics and Genotyping Facilities at Wellcome Sanger Institute and LabCorp (Laboratory Corporation of America) using support from 23andMe. FS, EV, MvD, BSTP and SEF are supported by the Max Planck Society. BSTP is supported by the Simons Foundation (Award ID: 514787). The authors gratefully acknowledge the ongoing contribution of the participants in the TEDS and their families. TEDS is supported by a programme grant to RP from the UK Medical Research Council (MR/M021475/1 and previously G0901245), with additional support from the US National Institutes of Health (AG046938). RP is supported by a Medical Research Council Professorship award (G19/2). The research leading to these results has also received funding from the European Research Council under the European Union’s Seventh Framework Programme (FP7/2007-2013)/ grant agreement n° 602768. This project has received funding from the European Union’s Horizon 2020 research and innovation programme, Marie Sklodowska Curie Actions (MSCA-ITN-2016) Innovative Training Networks (CAPICE grant 721567). KR is supported by a Sir Henry Wellcome Postdoctoral Fellowship.

