## Supplementary information for "Polygenic risk for psychiatric disorder reveals distinct association profiles across social behaviour in the general population"

**[Supplementary Methods](#_Toc74316503)**	[2](#_Toc74316503)

### **Supplementary Methods**

#### *Genome-wide summary statistics of psychiatric disorders*

ADHD-PCG/iPSYCH: We used GWAS summary statistics for clinical ADHD from a meta-analysis combining samples of the Psychiatric Genomic Consortium (PGC) and the Danish Lundbeck Foundation Initiative for Integrative Psychiatric Research (iPSYCH), comprising in total 20,183 ADHD cases and 35,191 controls^1^ (Supplementary Table 1). ADHD-iPSYCH is a case-control study (21.6% female ADHD-cases, and ~49% female controls) embedded within a nationwide population-based sample^1^. All participants were of European ancestry with an age range spanning infancy to adulthood^2^. ADHD cases (N=14,584) were identified via the Danish Psychiatric Central Research Register based on diagnoses by psychiatrists at a psychiatric hospital according to the International Classification of Diseases (ICD10, F90.0)^3^. Controls (N=22,492) were randomly selected from the same nationwide birth cohort and did not have a diagnosis of ADHD (F90.0) or moderate-severe intellectual disability (F71-F79)^1,2^. iPSYCH samples were derived from the Danish Newborn Screening Biobank hosted by Statens Serum Institute Genotypes were determined using the Illumina’s Beadarrays (PsychChip; Illumina, CA, San Diego, USA). Note that ADHD-iPSYCH controls are shared with the ASD-iPSYCH sample. ADHD-PGC comprises 7 case-control samples, and 4 family-based samples of predominantly white European ancestry including a total of 4,163 cases (~12-54% females) and 12,040 controls/pseudo-controls (~12-61% females), with ages spanning childhood to adulthood. Diagnoses of ADHD were based on the Diagnostic and Statistical Manual of Mental Disorders (DSM-III, DSM-IV, DSM-TR) or the ICD-10. Note, that ADHD-cases might have an additional diagnosis of ASD.

ASD-PGC/iPSYCH: GWAS summary statistics for clinical ASD were obtained from a meta-analysis combining samples from PGC and iPSYCH, with a total of 18,381 ASD cases and 27,969 controls^4^ (Supplementary Table 1). ASD-iPSYCH includes 13,076 cases and 22,664 controls, and is also based on the nationwide population-based iPSYCH sample. ASD cases (19-25.3% females) were identified via the Danish Psychiatric Central Research Register. ASD diagnoses were made by a psychiatrist in 2013 or earlier according to ICD-10 diagnoses of childhood autism, atypical autism, Asperger’s syndrome, other pervasive developmental disorders, and pervasive developmental disorder, unspecified (F84.0, F84.1, F84.5, F84.8, F84.9) with ages of first diagnoses ranging from 5 to 15 years. Controls (49.3% females) were randomly selected and did not have a diagnosis of ASD by 2013. ASD-PGC comprises 5 family-based cohorts of predominantly European ancestry^4,5^. ASD cases (N= 5,305) in PGC were identified based on research standard tools and expert clinical consensus diagnoses^4^. Individuals were excluded if assessments took place before the age of 36 months or if diagnostic criteria did not meet criteria of Autism Diagnostic Interview-Revised (ADI-R) or the Autism Diagnostic Observation Schedule (ADOS) domain scores^5^. Information on the male-female ratio was not available^6^.

BP-PGC: BP-PGC summary statistics are based on a meta-analysis of 32 cohorts with a total of 20,352 cases and 31,358 controls^7^ (Supplementary Table 1), all of European descent and aged 17 years or older^7^. There was no information on the male-female ratio^7^. BP diagnoses were made using structured diagnostic instruments from assessments by trained interviewers, clinician-administered checklists or medical record reviews, and were based on international consensus criteria (DSM-IV or ICD-10) for a lifetime diagnosis of BP^7^. In most cohorts, controls were screened for the absence of lifetime psychiatric disorders and randomly selected from the population^7^.

MD-PGC/UKBB: MD summary statistics are based on a meta-analysis of PGC and UK Biobank (UKBB) samples, with a total of 170,756 cases and 329,443 controls^8^ (Supplementary Table 1). MD-PGC includes 33 European ancestry cohorts with a total of 43,204 cases and 95,680 controls^9^. In most cohorts, cases were selected based on DSM-IV, ICD-10, or ICD-9, and did not have a diagnosis of BP, non-affective psychosis, MD related to substance use disorder, and mania or hypomania. Controls were predominantly selected using exclusion criteria with respect to a diagnosis of MD or depressive symptoms, BP or any other severe (mental) illnesses^9^. Information on participant age and a male-female ratio was not available^9,10^. The MD-UKBB sample is a UK population-based study recruited from the UK Biobank^8^ including 127,552 cases (65% females), and 233,763 controls (48% females) with an age range spanning from 39 to 73 years^10^. Cases were selected according to three depression phenotypes including broad depression (self-reported help-seeking behaviour for mental health difficulties from either a general practitioner or psychiatrist), probable MD^11^, and ICD-coded MD based on a primary or secondary diagnosis of MD in hospital admission records (ICD-9/10 codes F32, F33, F34, F38, F39 or non-cancer illness code 1286)^10^. Cases were excluded from the analysis if they were identified with bipolar disorder, schizophrenia, or personality disorder, and if there was evidence for being prescribed any antipsychotic medication^10^. Exclusion criteria from the control sample were a diagnosis of a depressive mood disorder from hospital admission records (ICD-9/10 codes F32, F33, F34, F38, and F39), a reported prescription for antidepressants or self-reported depression^10^ (Supplementary Table 1).

SCZ-PGC: Summary statistics are based on a meta-analysis of 46 ancestry-matched non-overlapping case-control samples and 3 family-based samples of European ancestry including a total of 33,640 cases and 43,456 controls/pseudo-controls^12^. Individuals with schizophrenia or schizoaffective disorder were included as cases, based on research-based assessment criteria or diagnoses by a clinician. A detailed cohort specific ratio of males is described elsewhere showing a balanced male/female representation in the majority of the cohorts^12^. Information on the age representation was not available^12^ (Supplementary Table 1).

In PGC and iPSYCH based studies, quality control, imputation, and association analyses were carried out for each cohort individually according to standard PGC settings using the PGC “ricopili” pipeline^12^. Genotypes were imputed against 1000 Genomes Project reference panels^13,14^ (Supplementary Table 1). Quality control criteria for the MD-UKBB sample are described in detail by Howard and colleagues^10^, and genotypes were imputed with IMPUTE4 against the HRC reference template^15,16^ (Supplementary Table 1).

#### *Description of ALSPAC*

Pregnant women resident in Avon, UK with expected dates of delivery 1st April 1991 to 31st December 1992 were invited to take part in the study^17,18^. The initial number of pregnancies enrolled is 14,541 (for these at least one questionnaire has been returned or a “Children in Focus” clinic had been attended by 19/07/99). Of these initial pregnancies, there was a total of 14,676 foetuses, resulting in 14,062 live births and 13,988 children who were alive at 1 year of age. When the oldest children were approximately 7 years of age, an attempt was made to bolster the initial sample with eligible cases who had failed to join the study originally. As a result, when considering variables collected from the age of seven onwards (and potentially abstracted from obstetric notes) there are data available for more than the 14,541 pregnancies mentioned above. The number of new pregnancies not in the initial sample (known as Phase I enrolment) that are currently represented on the built files and reflecting enrolment status at the age of 24 is 913 (456, 262 and 195 recruited during Phases II, III and IV respectively), resulting in an additional 913 children being enrolled. The phases of enrolment are described in more detail in the cohort profile paper and its update. The total sample size for analyses using any data collected after the age of seven is therefore 15,454 pregnancies, resulting in 15,589 foetuses. Of these 14,901 were alive at 1 year of age. A 10% sample of the ALSPAC cohort, known as the Children in Focus (CiF) group, attended clinics at the University of Bristol at various time intervals between 4 to 61 months of age. The CiF group were chosen at random from the last 6 months of ALSPAC births (1432 families attended at least one clinic). Excluded were those mothers who had moved out of the area or were lost to follow-up, and those partaking in another study of infant development in Avon. Please note that the study website contains details of all the data that are available through a fully searchable data dictionary and variable search tool.

#### *Social behavioural measures in the general population*

The SDQ is a brief behavioural screening questionnaire capturing behavioural symptoms^19^. The SDQ prosocial behaviour subscale consists of the items: (1) "Considerate of other people's feelings"; (2) "Shares readily with other children"; (3) "Helpful if someone is hurt"; (4) "Kind to younger children"; (5) "Often volunteers to help others". Each item was scored as either "not true" (2), "somewhat true" (1) or "certainly true" (0), and eventually added to a total sum score. The total sum of prosocial behaviour item scores was reverse-coded, such that higher scores reflect lower prosociality. The SDQ peer problem subscale includes the items (1) "Rather solitary, tends to play alone"; (2) "Has at least one good friend"; (3) "Generally liked by other children"; (4)"Picked on or bullied by other children"; 5)"Gets on better with adults than with other children". Items (4) and (5) were reverse-coded such that the total sum score of all peer problem items indicates increased peer-related problems.

A description of study numbers and demographics for each assessed social score in ALSPAC and TEDS are detailed in Table 1.

#### *Genome-wide genotyping information in ALSPAC and TEDS*

ALSPAC: Genome-wide genotyping in ALSPAC was performed using the Illumina HumanHap550 array^17,18^. Standard quality control was applied to the raw genome-wide data as described previously^20^. In brief, 8,981 subjects and 465,740 SNPs passed the quality control filters and were imputed to the Haplotype Reference Consortium (HRC) reference panel version r1.1 using the Sanger Imputation Server^15^. Only high-quality imputed SNPs (INFO > 0.8) with a genotype probability of > 0.8 in > 95% of the individuals, and a minor allele frequency (MAF) > 0.005 (*N*=7,337,399) were included in subsequent analyses.

TEDS: Genome-wide genotyping data were assessed using the Affymetrix GeneChip 6.0 or the Illumina HumanOmniExpressExome-8v1.2 arrays. Typical quality control procedures were followed and have been described in detail elsewhere^21^. In short, quality-controlled genotypes from the two platforms (7,289 individuals and 559,772 SNPs genotyped on Illumina and 3,057 individuals and 635,269 SNPs genotyped on Affymetrix) were imputed separately to the HRC reference panel version r1.1^15^, and then harmonised. Only high-quality imputed SNPs (INFO>0.75, genotyping-missingness<0.02, individual-missingness<0.02, MAF>0.005, and Hardy Weinberg equilibrium *P*-value>0.00001; *N*=7,363,646) were included for further analyses. The following exclusions were applied: extreme perinatal conditions and severe medical conditions.

#### *SNP-h^2^ analyses in ALSPAC using genome-wide summary statistics*

For SNP-heritability (SNP-h^2^) analyses of social behaviour in ALSPAC children, we conducted genome-wide association studies (GWAS) using untransformed social scores for both low prosociality (7, 10, 13, 14, and 17 years of age), and peer problems (7, 10, 13, 14, and 17 years of age). Avoiding data transformation, we use a count data-based regression approach to either regress positively skewed peer-problems or low-prosociality scores on allele dosage, age, sex, and the first two most significant ancestry-informative principal components (PC; correcting for subtle differences in population structure).

We apply a quasi-Poisson model that accommodates count data with a variance exceeding the mean (over-dispersion, Table 1).

The quasi-Poisson regression was model given by

$$\log(\mu)=\beta_{0}+ \beta_{{SNP}_{i}}x_{{SNP}_{ij}}+\beta_{age}x_{{age}_{j}}+\beta_{sex}x_{{sex}_{j}}+ \beta_{PC1}x_{{PC1}_{j}}+\beta_{PC2}x_{{PC2}_{j}}+\varepsilon_{j}$$

$$var\left( y \right)=\Phi\mu$$

where log changes in $\mu$, the mean value of a social trait, are a function of $x_{{SNP}_{ij}}$ the allele dosage for the *i*-th high-quality imputed SNP for the *j-*th participant, $x_{{age}_{j}}$the age of assessment, $x_{{sex}_{j}}$ the dummy coded sex with 1 indicating male and 0 female, and $x_{{PC1}_{j}}$ and $x_{{PC2}_{j}}$ the first two principal components for the *j-*th participant. The quasi-Poisson model accounts for over-dispersion in count data by defining the variance of y by the mean $\mu$ proportional to its dispersion parameter $\Phi$, where $\Phi$ is given by Pearson’s $X^{2}$ divided by the residual degrees of freedom (df): $\hat{\Phi}=\frac{X^{2}}{df}$ with $\hat{\Phi}$> 1 indicating over-dispersion^22^. Note, that a quasi-Poisson regression does not model an error distribution, with the deviance being identical to the one derived from a Poisson distribution. Therefore, model fit cannot be assessed using a likelihood-ratio test^23^. However, compared to the conceptually related negative binomial model, the quasi-Poisson model is easier to integrate within a genome-wide analysis framework, due to less restrictive modelling assumptions^24^. We demonstrate an improved model fit using count data approaches, allowing for over-dispersion, by comparing the model fit of a negative binomial model with an ordinary least squares model (Supplementary Table 4). After creating genome-wide genotype dosage files in hdf5 file format, using the HASE framework^25^, we applied quasi-Poisson regressions using custom-based R scripts, as count models are not yet implemented in standard GWAS software.

SNP-h^2^ for social behaviour (low-prosociality and peer-problem scores) and psychiatric conditions were estimated from GWAS summary statistics using unconstrained linkage disequilibrium score (LDSC) regression^26^ to inform power analyses.

SNP-h^2^ estimates describe here either the proportion of phenotypic variance (social behaviour) or liability to disorder, as tagged by common SNPs on genotyping arrays (Supplementary Tables 7 and 8). In LDSC, SNP-h^2^ estimates represent the slope of the regression of genome-wide Χ^2^-statistics against the corresponding LD scores, while the intercept minus one estimates the mean contribution of confounding bias due to the inflation in the mean Χ^2^-statistic^26^. For completeness, we also calculated genomic control λ (λ_GC_) estimates which assess the extent of inflation in GWAS test statistics due to population stratification^27^. SNP-h^2^ for psychiatric disorders was estimated on the liability scale, assuming a population prevalence of 0.05 for ADHD^28^, 0.012 for ASD^4^, 0.006 for BP^29^, 0.162 for MD^30^, and 0.007 for schizophrenia^31^.

All analyses were performed with LDSC software using pre-computed LD scores based on European-ancestry samples of the 1000 Genomes European Project^26^.

All SNP-h^2^ analyses were corrected for multiple testing accounting for 14 different social-behavioural scores: 7 low-prosociality scores (parent reports at 7, 10, 13, 14, and 17 years of age, and teacher reports at 8 and 11 years of age) and 7 peer-problem scores (parent reports at 7, 10, 13, 14, and 17 years of age, and teacher reports at 8 and 11 years of age) in ALSPAC.

#### *Univariate polygenic scoring analyses*

Polygenic risk scores (PRS) for psychiatric disorder were constructed for ALSPAC and TEDS participants using PLINK software^33^. Consistent with current guidelines^34^, clinical summary statistics from ADHD-PGC/iPSYCH, ASD-PGC/iPSYCH, BP-PGC, MD-PGC/UKBB, and schizophrenia-PGC were clumped (LD-r2>0.25, ±500 kb) using PLINK software^33^. Risk variants were selected from summary statistics across a range of P-value thresholds (0.001≤P_T_<1). In ALSPAC, PRS were constructed for unrelated ALSPAC children and adolescents (genomic relatedness<0.125), based on high-quality imputed SNPs (INFO>0.8, 95%-posterior genotyping probability>0.9, MAF>0.005). In TEDS, PRS were generated for dizygotic twin pairs and a single twin from each monozygotic twin pair, using high-quality imputed SNPs in TEDS (INFO>0.75, genotyping-missingness<0.02, individual-missingness<0.02, MAF>0.005, and Hardy Weinberg equilibrium *P*-value>0.00001). The log odds of genetic SNP effects were aligned to indicate alleles with increased risk for psychiatric disorder, and PRS were Z-standardised.

To estimate the association between polygenic risk for psychiatric disorder and social behaviour in ALSPAC we applied a negative binomial regression where log changes in the mean $\mu$ are given by

$$\log\left( \mu\right)=\beta_{0}+ \beta_{PRS}x_{{PRS}_{j}}+\beta_{age}x_{{age}_{j}}+\beta_{sex}x_{{sex}_{j}}+ \beta_{PC1}x_{{PC1}_{j}}+\beta_{PC2}x_{{PC2}_{j}}+\varepsilon_{j}$$

$$var\left( y \right)=\mu+\mu^{2}/\Phi$$

with $x_{{PGS}_{j}}$being the polygenic risk score, $x_{{age}_{j}}$ the age of assessment, $x_{{sex}_{j}}$ the dummy coded sex with 1 indicating male and 0 female, and $x_{{PC1}_{j}}$ and $x_{{PC2}_{j}}$ the first two principal components for the *j*-th participant. Here, the variance of y is equal to the mean adjusted by the inverse of the overdispersion dispersion parameter $\Phi$, scaled by the square of the mean.

The negative binomial model accounts for an over-dispersed error distribution in count data. The model can be fitted using a maximum likelihood approach with derived parameters following a defined probability distribution^23^. Thus, models can be compared using likelihood-ratio test and the strength of the association can be assessed using quasi-R^2^ measures.

For analyses in TEDS, we implemented a generalized linear mixed-effects modelling approach that additionally allowed for a random intercept $u_{i}$accounting for within family relatedness:

$\log\left( \mu\right)=\beta_{0}+ \beta_{PRS}x_{{PRS}_{j}}+\beta_{age}x_{{age}_{j}}+\beta_{sex}x_{{sex}_{j}}+ \beta_{PC1}x_{{PC1}_{j}}+\beta_{PC2}x_{{PC2}_{j}}+\varepsilon_{j}+ u_{i}$.

As a measure to assess the improvement in model fit attributable to $\beta_{PRS}$ as a predictor, we assessed ΔMc Fadden’s R^2^ which was given by

$$\Delta{Mc Fadden^{'}s R}^{2}=\left( 1- \frac{LL\left( Full model \right)}{LL\left( Intercept model \right)} \right)- \left( 1- \frac{LL\left( Baseline model \right)}{LL\left( Intercept model \right)} \right)$$

as the difference between one minus the proportion of the log likelihood (LL) of the full model representing the regression function including $\beta_{PRS}$ as a predictor against the LL of the intercept model, and one minus the proportion of the LL of the baseline model representing the regression function excluding $\beta_{PRS}$ as a predictor against the LL of the intercept model.

#### *Power of univariate PRS analyses in ALSPAC*

We assessed the power to detect univariate polygenic trait-disorder associations in ALSPAC using the R software package avengeme^35^. Power estimates were informed by the estimated SNP-h^2^ of the psychiatric disorder, based on summary statistics, the sample sizes of discovery and target samples, and the number of genetic variants used to construct PRS (Table 1, Supplementary Tables 1,7). We set the proportions of SNPs with no causal effect on the discovery sample (i.e. the presence of large single genetic effects, pi0) to either pi0=0, or 0.95. We estimated power under two covariance settings across a range of applied *P*-value selection thresholds (0.001≤*P*_T_<1): (1) a fixed trait-disorder covariance governed by the SNP-h^2^ of the psychiatric condition and (2) the estimated genetic trait-disorder covariance as observed in the univariate regression model. Note, that a drop in power of the latter approach compared to condition 1, reflects the lack in observed genetic covariance, and is, thus, not governed by a potential lack in SNP-h².

#### *Random-effects meta-regression*

In order to investigate variation in estimates in the association between polygenic risk for psychiatric disorder and social traits (PRS effects), we conducted 5 random-effect meta-analyses (R:metafor). Specifically, we combined for each disorder the 14 ALSPAC-based and 15 TEDS-based PRS effects for SDQ-based social scores (β_i_) to assess age-, rater- and trait-specific differences in polygenic associations. First, we tested a model that was given by

$$\beta_{i}=\theta_{0}+\theta_{age}x_{{age}_{i}}+ \theta_{rater}x_{{rater}_{i}}+\theta_{trait}x_{{trait}_{i}}+\theta_{cohort}x_{{cohort}_{i}} + u_{ij}$$

including a random intercept ($u_{i}$), and fixed effects for $x_{{cohort}_{i}}$ the dummy-coded indicator of assessment within ALSPAC (0) versus TEDS (1), $x_{{age}_{i}}$ the median age within the *i*-th social score, $x_{{rater}_{i}}$ the dummy-coded parent (0) versus teacher-report (1), and $x_{{trait}_{i}}$the dummy-coded low-prosociality (0) versus peer-problems (1) score assuming that $u_{i}\sim N(0,\tau^{2})$. $\tau^{2}$ denotes the amount of residual heterogeneity among the true effects^36^. Note that peer problems and low prosociality were each assessed with a 5-item subscale of the SDQ and are recorded as item counts. We first established whether cohort-specific fixed effects contributed to the model. Next, we dropped the fixed effect for cohort, and identified for each disorder the most parsimonious model, confirmed by likelihood-ratio tests at *P*>0.05 against a full model that included fixed effects for age, reporter, and trait, and applied Cochran’s Q test to assess residual heterogeneity.

We accounted for interrelatedness among PRS effects within each cohort using an approach that is analogous to models that account for correlated phylogenetic histories with the variance covariance matrix^37^. The variance covariance matrix was given by $V =SV_{pheno}S$ with $S$ being a diagonal matrix with the standard errors (SE) of PRS effect estimates

$S=\left[ \begin{matrix} {SE}_{PRS 1} & & \\ & {SE}_{PRS 1} & \\ & & \begin{matrix} \ddots& \\ & {SE}_{PRS 14} \end{matrix} \end{matrix} \right]$

and $V_{pheno}$ being the Pearson’s phenotypic correlation matrix of the social scores:

$V_{pheno}=\left[ \begin{aligned} \begin{matrix} 1 & {corr}_{2 1} & \cdots\\ {corr}_{1 2} & 1 & \\ \vdots& \vdots& \ddots\end{matrix} \\ \begin{matrix} {corr}_{1 14} & {corr}_{2 14} & \ldots\end{matrix} \end{aligned}\begin{matrix} {corr}_{14 1} \\ \\ \begin{matrix} \vdots\\ 1 \end{matrix} \end{matrix} \right]$.

#### *Web resources*

ALSPAC data dictionary: http://www.bris.ac.uk/alspac/researchers/data-access/data-dictionary/

ALSPAC variable catalogue: http://www.bristol.ac.uk/alspac/researchers/access/

PGC: http://www.med.unc.edu/pgc

iPSYCH: http://ipsych.au.dk

UKBB: https://www.ukbiobank.ac.uk/

PLINK: https://www.cog-genomics.org/plink2

HRC: http://www.haplotype-reference-consortium.org/

SANGER IMPUTATION SERVER: https://imputation.sanger.ac.uk/

LDSC: https://github.com/bulik/ldsc

R: https://www.r-project.org/

METAFOR: http://www.metafor-project.org/doku.php

### **Supplementary Figures**

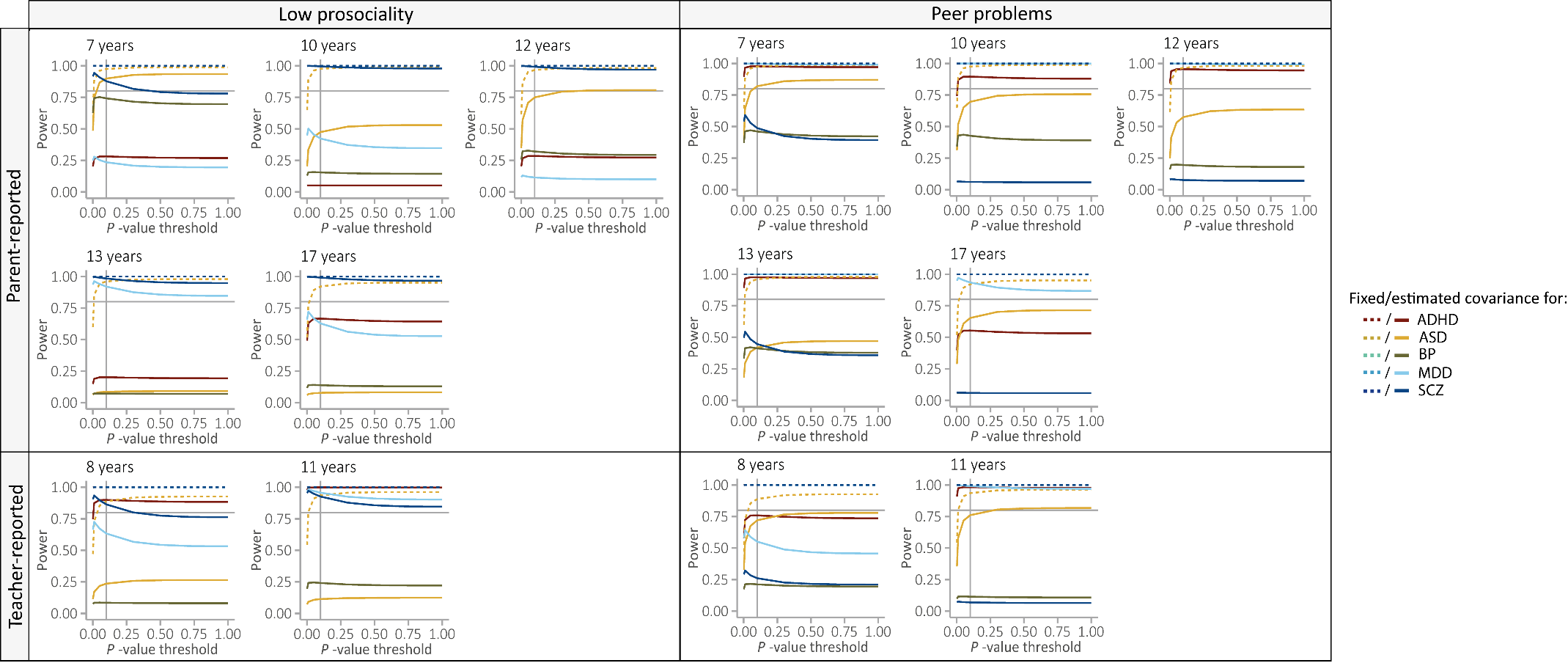

**Supplementary Figure 1:** Power-analysis of psychiatric PRS constructed for different P-value thresholds in the ALSPAC discovery samples for low-prosociality and peer-problem scores reported by parents or teachers at the age of 7 to 17 years.

Analyses were conducted using the R software package avengeme^35^ accounting for the heritability of the discovery sample, the sample sizes of the discovery and the target sample, and the number of genetic variants used to construct PRS. Here, the proportion of markers with no effect on the training set (pi0) was set to 0.95.

ADHD - Attention-deficit/hyperactivity disorder; ALSPAC - Avon Longitudinal study of Parents and Children; ASD - Autism spectrum disorders; BP - Bipolar disorder; MD- Major depression; PRS-Polygenic risk scores; SCZ – Schizophrenia

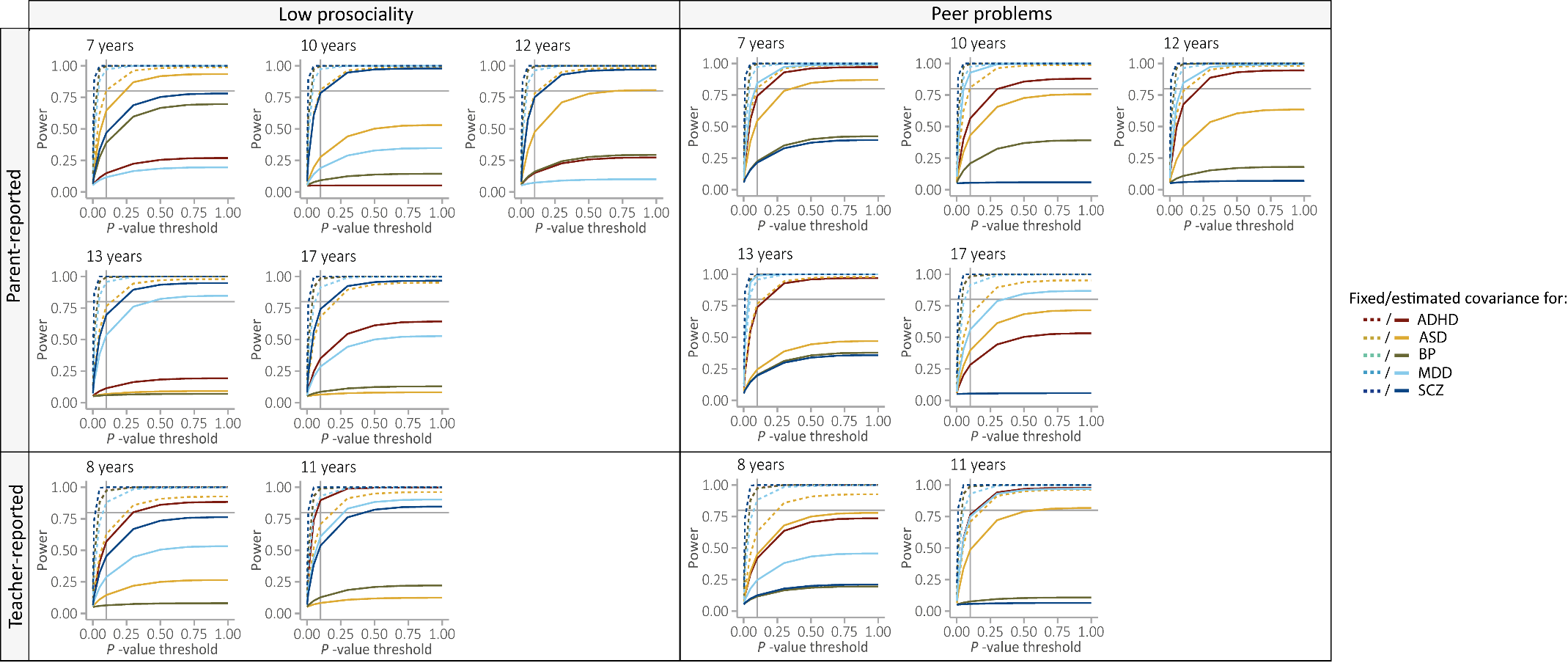

**Supplementary Figure 2:** Power-analysis of psychiatric PRS constructed for different P-value thresholds in the ALSPAC discovery samples for low-prosociality and peer-problem scores reported by parents or teachers at the age of 7 to 17 years.

Analyses were conducted using the R software package avengeme^35^ accounting for the heritability of the discovery sample, the sample sizes of the discovery and the target sample, and the number of genetic variants used to construct PRS. Here, the proportion of markers with no effect on the training set (pi0) was set to 0.

ADHD - Attention-deficit/hyperactivity disorder; ALSPAC - Avon Longitudinal study of Parents and Children; ASD - Autism spectrum disorders; BP - Bipolar disorder; MD- Major depression; PRS-Polygenic risk scores; SCZ – Schizophrenia

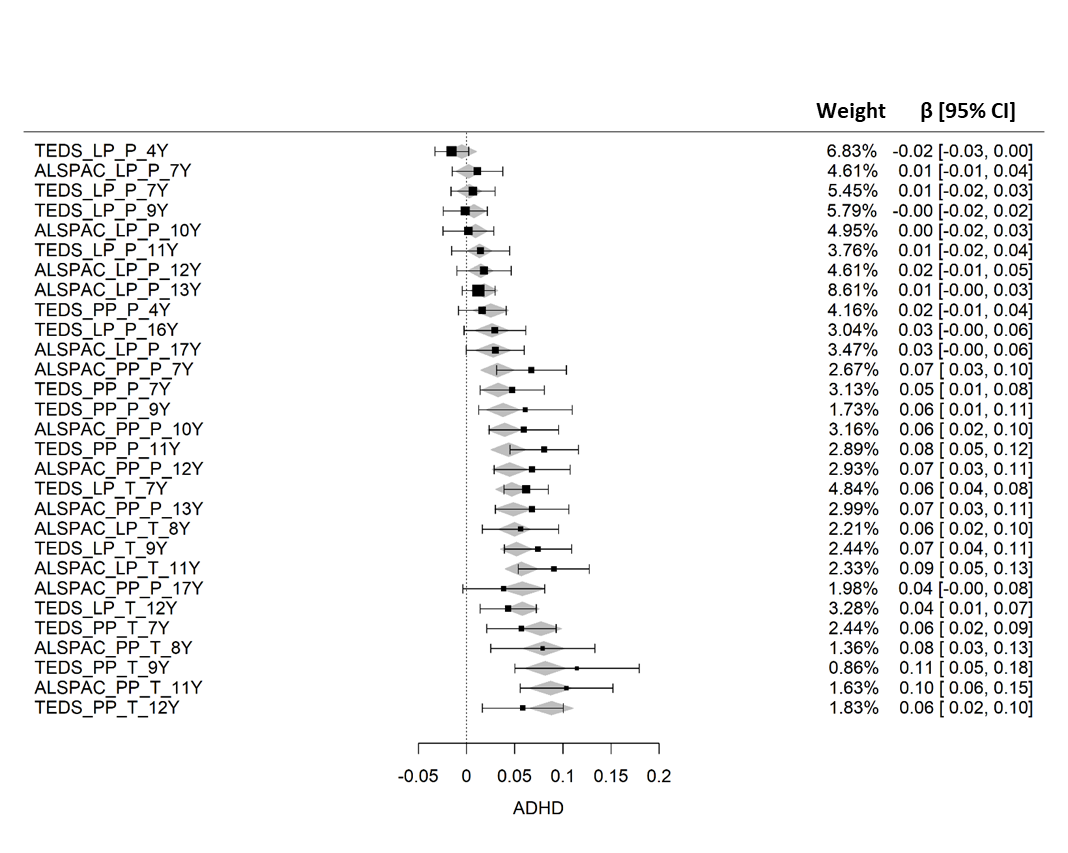

**Supplementary Figure 3:** Forest plot showing both univariate ADHD-PRS effects (β) on low-prosocialty and peer-problem scores (black squares) and predicted ADHD-PRS effects ($\hat{\beta}$; grey diamonds) using the most-parsimonious random-effects meta-regression model with respective 95% CI bands.

ADHD-PRS association effects (based on negative binominal regression) were combined across 29 social symptoms (14 ALSPAC-based + 15 TEDS-based; at P_T_ ≤ 0.1) using random-effects meta-regression, accounting for phenotypic correlations between social scores (weights). The most-parsimonious predictors for heterogeneity in ADHD-PRS effects included age (years), reporter (parent vs teacher), and trait (low prosociality vs peer problems).

Low-prosociality and peer-problem scores were assessed using the Strengths-and-Difficulties questionnaire.

ADHD - Attention-deficit/hyperactivity disorder; ALSPAC - Avon Longitudinal study of Parents and Children; CI - Confidence interval; LP - Low prosociality; P- Parent-report; PP - Peer problems; PRS-Polygenic risk scores; T- Teacher-report; TEDS - Twins Early Development Study; Y – Age in years

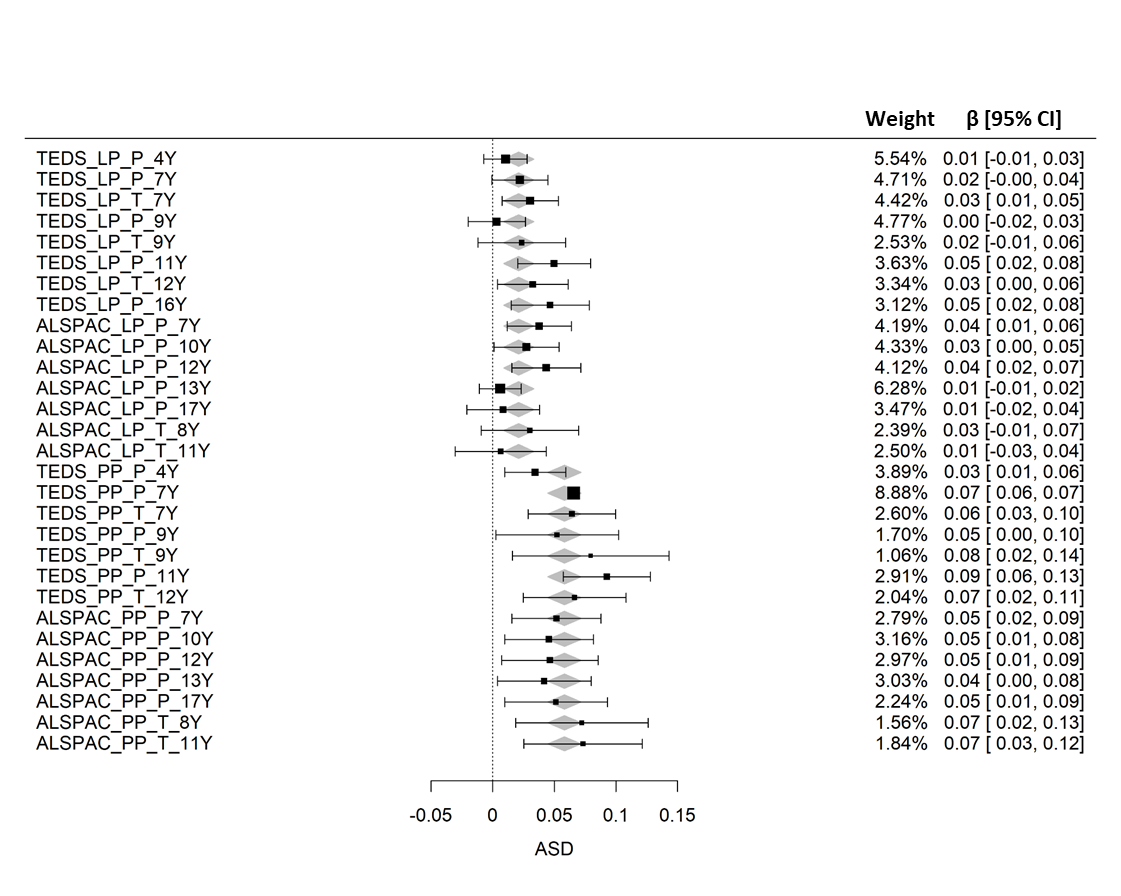

**Supplementary Figure 4:** Forest plot showing both univariate ASD-PRS effects (β) on low-prosocialty and peer-problem scores (black squares) and predicted ASD-PRS effects ($\hat{\beta}$; grey diamonds) using the most-parsimonious random-effects meta-regression model with respective 95% CI bands.

ASD-PRS association effects (based on negative binominal regression) were combined across 29 social symptoms (14 ALSPAC-based + 15 TEDS-based; at P_T_ ≤ 0.1) using random-effects meta-regression, accounting for phenotypic correlations between social scores (weights). The most-parsimonious predictor for heterogeneity in ASD-PRS effects included trait (low prosociality vs peer problems).

Low-prosociality and peer-problem scores were assessed using the Strengths-and-Difficulties questionnaire.

ASD - Autism spectrum disorders; ALSPAC - Avon Longitudinal study of Parents and Children; CI - Confidence interval; LP - Low prosociality; P- Parent-report; PP - Peer problems; PRS-Polygenic risk scores; T- Teacher-report; TEDS - Twins Early Development Study; Y – Age in years

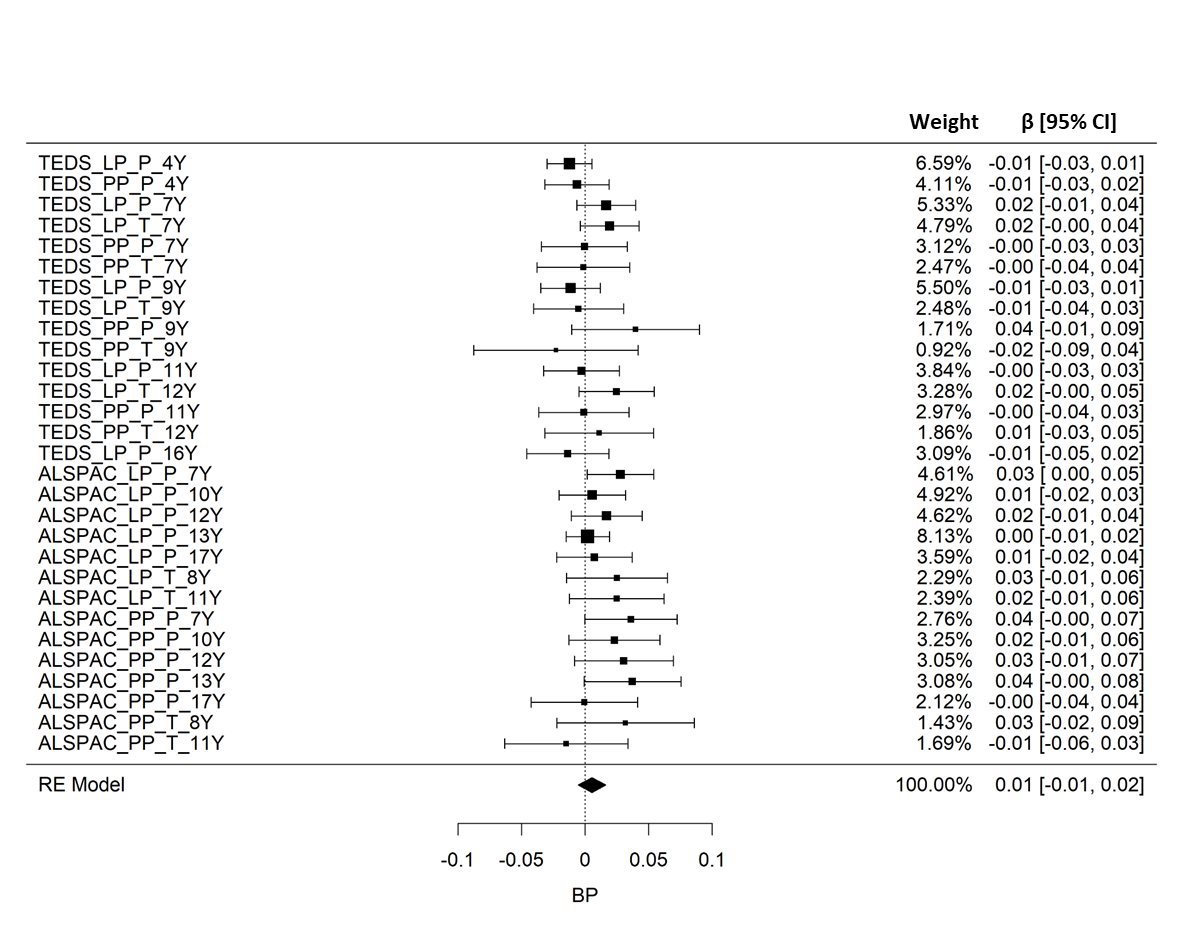

**Supplementary Figure 5:** Forest plot showing both univariate BP-PRS effects (β) on low-prosocialty and peer-problem scores (black squares) and predicted BP-PRS effect ($\hat{\beta}$; black diamond) using the most-parsimonious random-effects meta-regression model with respective 95% CI bands.

BP-PRS association effects (based on negative binominal regression) were combined across 29 social symptoms (14 ALSPAC-based + 15 TEDS-based; at P_T_ ≤ 0.1) using random-effects meta-regression, accounting for phenotypic correlations between social scores (weights). The most-parsimonious model for BP-PRS effects was the intercept model.

Low-prosociality and peer-problem scores were assessed using the Strengths-and-Difficulties questionnaire.

ALSPAC - Avon Longitudinal study of Parents and Children; BP - Bipolar disorder; CI - Confidence interval; LP - Low prosociality; P- Parent-report; PP - Peer problems; PRS-Polygenic risk scores; T- Teacher-report; TEDS - Twins Early Development Study; Y – Age in years

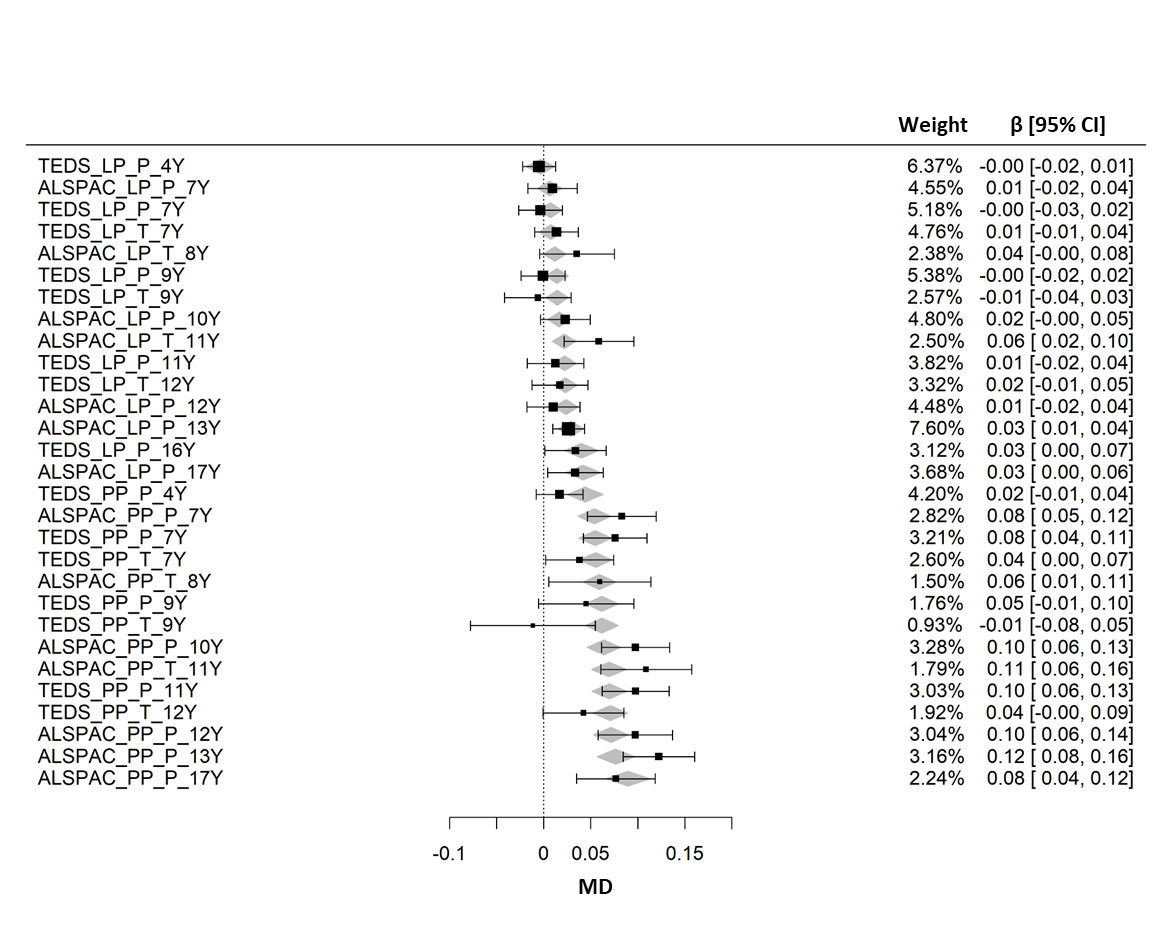

**Supplementary Figure 6:** Forest plot showing both univariate MD-PRS effects (β) on low-prosocialty and peer-problem scores (black squares) and predicted MD-PRS effects ($\hat{\beta}$; grey diamonds) using the most-parsimonious random-effects meta-regression model with respective 95% CI bands.

MD-PRS association effects (based on negative binominal regression) were combined across 29 social symptoms (14 ALSPAC-based + 15 TEDS-based; at P_T_ ≤ 0.1) using random-effects meta-regression, accounting for phenotypic correlations between social scores (weights). The most-parsimonious predictor for heterogeneity in MD-PRS effects included age (years) and trait (low prosociality vs peer problems).

Low-prosociality and peer-problem scores were assessed using the Strengths-and-Difficulties questionnaire.

ALSPAC - Avon Longitudinal study of Parents and Children; CI - Confidence interval; LP - Low prosociality; MD- Major depression; P- Parent-report; PP - Peer problems; PRS-Polygenic risk scores; T- Teacher-report; TEDS - Twins Early Development Study; Y – Age in years

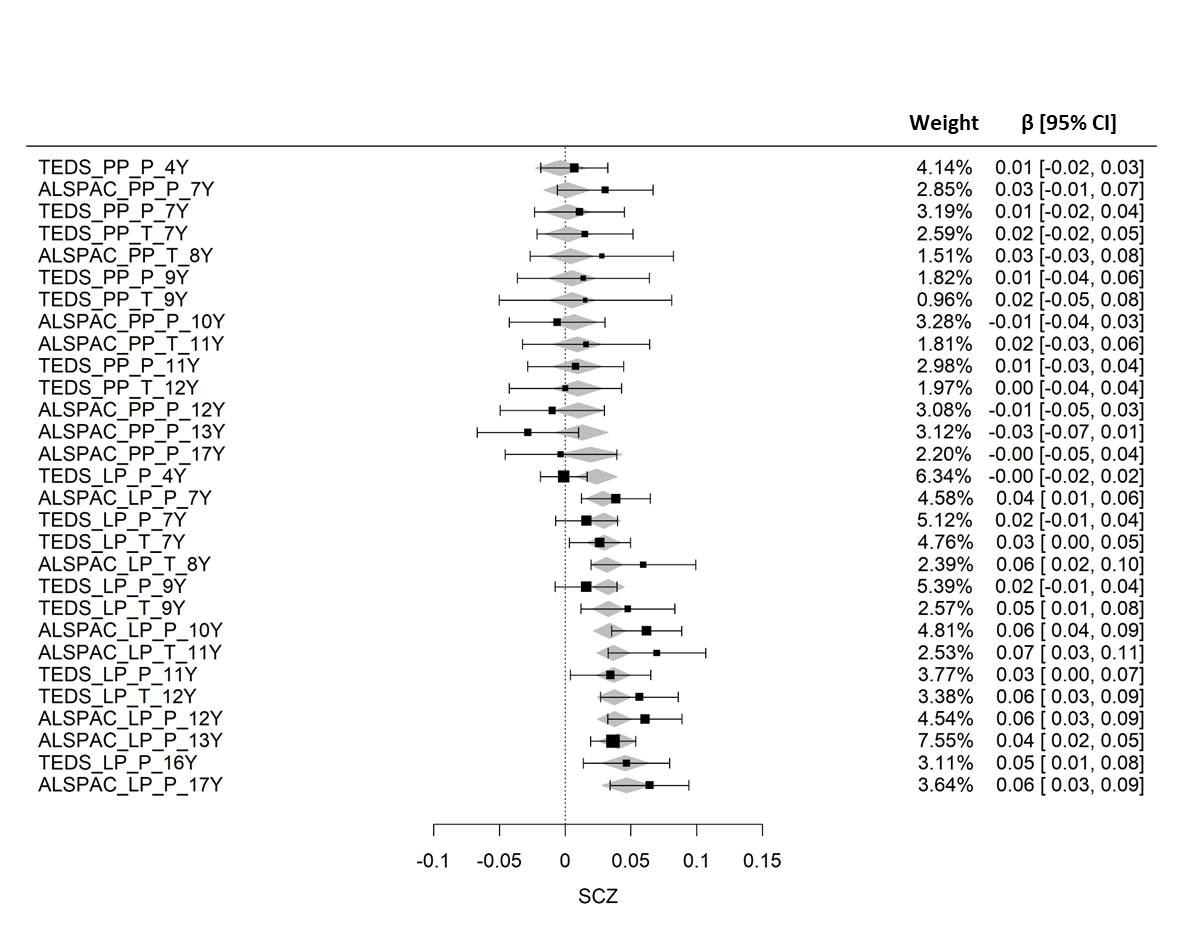

**Supplementary Figure 7:** Forest plot showing both univariate schizophrenia-PRS effects (β) on low-prosocialty and peer-problem scores (black squares) and predicted schizophrenia -PRS effects ($\hat{\beta}$; grey diamonds) using the most-parsimonious random-effects meta-regression model with respective 95% CI bands.

Schizophrenia -PRS association effects (based on negative binominal regression) were combined across 29 social symptoms (14 ALSPAC-based + 15 TEDS-based; at P_T_ ≤ 0.1) using random-effects meta-regression, accounting for phenotypic correlations between social scores (weights). The most-parsimonious predictor for heterogeneity in schizophrenia -PRS effects included age (years) and trait (low prosociality vs peer problems).

Low-prosociality and peer-problem scores were assessed using the Strengths-and-Difficulties questionnaire.

ALSPAC - Avon Longitudinal study of Parents and Children; CI - Confidence interval; LP - Low prosociality; P- Parent-report; PP - Peer problems; PRS-Polygenic risk scores; SCZ- Schizophrenia; T- Teacher-report; TEDS - Twins Early Development Study; Y – Age in years
